# Fibromuscular Dysplasia in Acute Type A Aortic Dissection: A Common yet Underrecognized Vascular Phenotype in Women

**DOI:** 10.64898/2026.11.03.26367049

**Authors:** Annick C. Weustink, Adine R. de Keijzer, Ricardo P. J. Budde, Jorie Versmissen, Tjebbe W. Galema, Kevin M. Veen, Johanna J. M. Takkenberg, Bardia Arabkhani, Adriaan Coenen

## Abstract

**Objectives:** Fibromuscular dysplasia (FMD) is a non-inflammatory, non-atherosclerotic arterial disease characterized by multifocal arterial beading that predominantly affects women. Although FMD commonly involves the renal and carotid arteries, its relationship with acute type A aortic dissection (ATAAD) remains poorly understood. We investigated the proportion of patients with FMD among patients with ATAAD, focusing on sex-based differences.

**Materials and Methods:** Patients aged ≤70 years with ATAAD and ECG-triggered aortic CT angiography (CTA) were consecutively included in this retrospective single-center study. FMD was defined as multifocal arterial beading (“string-of-beads” appearance) in at least one vascular territory, according to international consensus criteria. The highest-quality CTA examination was evaluated by an experienced cardiovascular radiologist. All suspected FMD cases were independently reviewed by a second experienced cardiovascular radiologist, with the final diagnosis established by consensus. Proportions were reported with 95% confidence intervals and compared between sexes.

**Results:** Of 214 patients screened, 200 were included (138 men and 62 women). FMD was confirmed in 18/200 patients (9%; 95% CI, 6–14%), none of whom had a prior FMD diagnosis. FMD was more frequent in women (12/62, 19%; 95% CI, 11–31%) than men (6/138, 4%; 95% CI, 2–9%; P<0.001). FMD predominantly involved the renal arteries. Additional carotid CTA was available in 49/200 patients (25%) and in 7/18 patients with FMD (39%); three of these seven patients had carotid FMD.

**Conclusion:** FMD was observed in a substantial proportion of patients presenting with ATAAD, particularly women. These findings suggest that FMD is an underrecognized vascular phenotype in patients with ATAAD.

**Key Points:** *Question:* The proportion of patients with fibromuscular dysplasia among those presenting with acute type A aortic dissection remains poorly understood, particularly regarding sex-based differences.

*Findings:* Fibromuscular dysplasia was identified on CT angiography in 19% of women and 4% of men with acute type A aortic dissection (9% overall).

*Relevance Statement:* Fibromuscular dysplasia represents an underrecognized vascular phenotype in patients with acute type A aortic dissection, particularly women. Its coexistence with connective tissue disease highlights the heterogeneity of this patient population.

## Introduction

Acute type A aortic dissection (ATAAD) is a life-threatening cardiovascular emergency. The global incidence of aortic dissection is estimated at 3–6 cases per 100,000 person-years, with ATAAD accounting for nearly two-thirds of cases.[1; 2] Despite advances in diagnostic and surgical techniques, ATAAD continues to carry a 30-day mortality exceeding 20%.[3] Prompt surgical repair remains the standard of care; however, outcomes are influenced by patient age, hemodynamic status, and institutional experience.[4–6]

Sex differences in presentation and outcomes have been increasingly recognized. Women are typically older at ATAAD presentation and are more likely to exhibit atypical symptoms.[7]

Fibromuscular dysplasia (FMD) is a non-inflammatory, non-atherosclerotic disease affecting arterial vessel walls. Arterial manifestations include beading, stenosis, aneurysms, tortuosity, and dissections. The most common subtype of FMD presents with a “string-of-beads” appearance. FMD most commonly affects the renal and carotid arteries, but other arteries can also be affected.[8; 9] Clinical manifestations of FMD include hypertension, headache, and pulsatile tinnitus, but FMD can also be asymptomatic.[10]

The exact prevalence of FMD remains uncertain. However, data from potential kidney donors undergoing CTA suggest an overall prevalence of 2.6–2.8%, with a higher prevalence among women (approximately 3.9%) than among men (1.2%).[11; 12] When invasive angiography was used to detect FMD in potential kidney donors, a prevalence of 4.4% was reported.[13] However, these prevalence estimates may be influenced by selection bias, because potential kidney donors are typically younger, healthier, and largely asymptomatic compared with patients with ATAAD.[11] Hendricks et al. showed that in an older population with hypertension and a higher burden of atherosclerosis, the prevalence of FMD was 5.8%, higher than the 2.3% prevalence observed in the potential kidney donor comparison group in that study.[14]

Although traditionally considered a disease mainly affecting medium-sized vessels, FMD has recently been associated with changes in the thoracic aorta. A cohort study reported that patients with FMD had slightly larger diameters of the ascending aorta compared with age- and sex-matched controls.[15] Case reports have described ATAAD in patients with FMD.[16; 17] However, the proportion of patients with ATAAD who have FMD has not been systematically investigated, and potential sex-based differences remain unknown.

Therefore, the aim of this cross-sectional analysis was to determine the proportion of FMD among patients aged ≤70 years with ATAAD and to evaluate sex-based differences.

## Methods

The study was approved by the institutional review board of Erasmus MC, which waived the requirement for informed consent because of the retrospective nature of the study. The study was conducted in accordance with the Declaration of Helsinki and applicable institutional and national regulations.

We conducted a retrospective single-center cohort study including patients aged 18–70 years who were surgically treated for ATAAD at Erasmus Medical Center (EMC) between 2007 and 2024. We restricted the analysis to patients ≤70 years of age because we hypothesized that the association between FMD and ATAAD would be more apparent in a younger population, in whom competing vascular pathologies such as atherosclerosis are less prevalent.

Patients were eligible for inclusion if they had a confirmed diagnosis of ATAAD and an available diagnostic ECG-triggered CT angiography (CTA) of the entire aorta. When multiple CTA examinations were available, the examination with the highest image quality was selected for analysis.

All scans were reviewed by a radiologist (AC, with 12 years of experience in cardiovascular imaging). Cases suspected of FMD were independently reviewed by a second radiologist (AW, with 20 years of experience in cardiovascular imaging), and the final diagnosis was established by consensus. All available medium-sized vessels were evaluated for FMD (renal, carotid, vertebral, subclavian, coronary, celiac, mesenteric, and iliac arteries). When additional CTA examinations of the carotid arteries were available, these were also reviewed for the presence of FMD.

Radiological signs of FMD were assessed according to the diagnostic criteria outlined in the First International Consensus on the Diagnosis and Management of Fibromuscular Dysplasia. Definite FMD was defined as multifocal beading (“string-of-beads” appearance) in at least one vessel.[8] Consistent with the international consensus criteria, isolated focal stenoses or aneurysms were not considered diagnostic of FMD.

Baseline clinical characteristics were obtained from our institution’s extended cohort of the SIZE MATTERS study, a multicenter project investigating sex-specific characteristics and outcomes in thoracic aortic disease. These characteristics included demographics, cardiovascular risk factors, and information on genetically confirmed or suspected connective tissue disease.[18]

Continuous variables are presented using mean ± standard deviation, and categorical variables are presented as counts and percentages. The proportion of patients with FMD is reported with two-sided 95% confidence intervals calculated using the Wilson score method. Differences in continuous variables were assessed using the independent-samples t-test. Differences in categorical variables were assessed using the χ² test; Fisher’s exact test was used when expected cell counts were less than five. A P-value <0.05 was considered statistically significant. Data were analyzed using IBM SPSS Statistics for Windows (version 28.0; IBM Corp., Armonk, NY, USA).

## Results

### Study population and FMD detection

In total, 214 patients aged 18–70 years underwent emergency surgery for ATAAD at our institution between 2007 and 2024. Fourteen patients were excluded due to the absence of diagnostic CTA imaging. Ultimately, 200 patients with ATAAD were included, of whom 62 (31%) were female. All CTA examinations were available with a reconstructed slice thickness of ≤1 mm. Initial screening of the CTA studies identified 22 patients with suspected FMD. After consensus review by two experienced radiologists, the diagnosis of FMD was confirmed in 18/200 patients (9.0%; 95% CI, 5.8–13.8%), whereas in four patients the initially suspected multifocal beading was considered insufficiently convincing for a diagnosis of FMD (Figure 1). None of the 18 patients with FMD had a known diagnosis of FMD prior to presentation with ATAAD.

**Figure 1:**
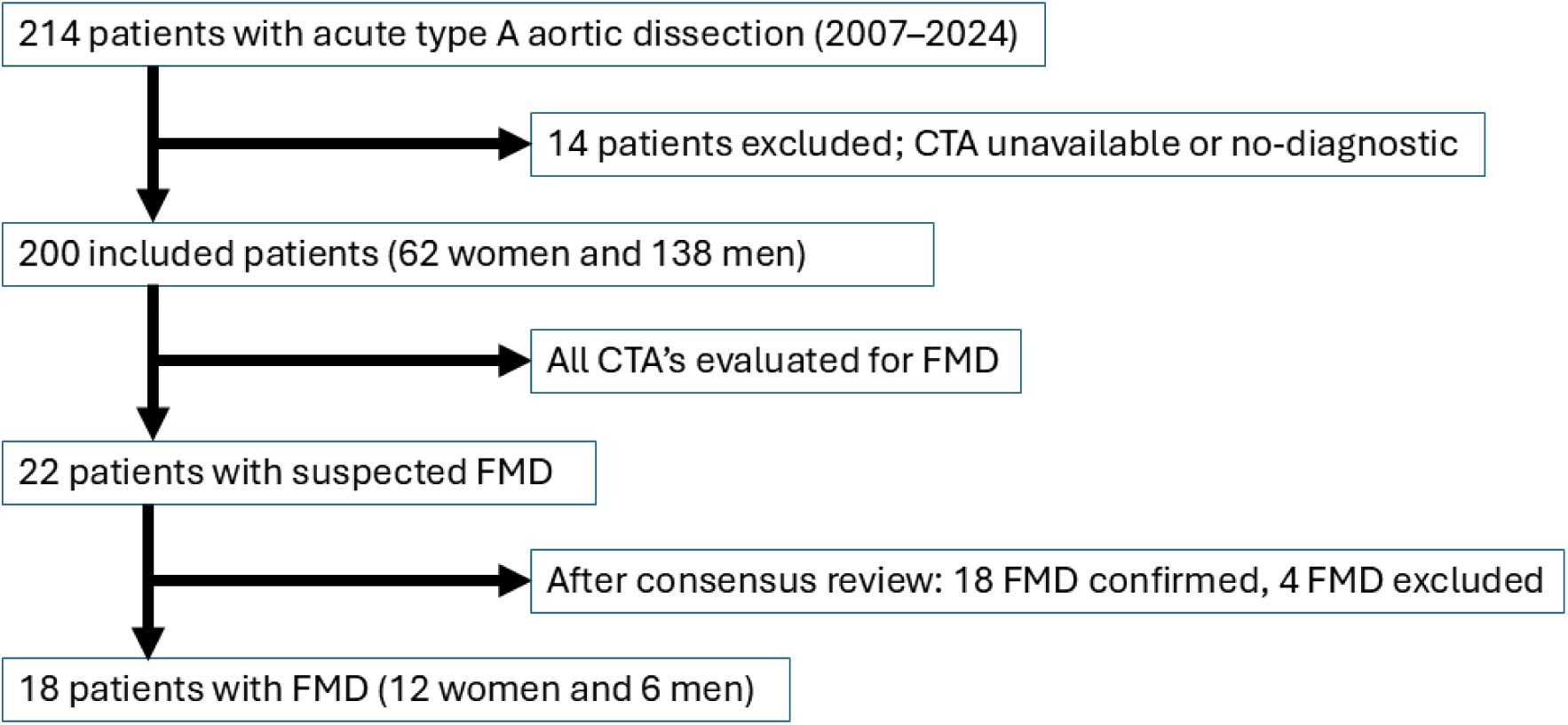
Study population flowchart. Flowchart illustrating patient selection and classification of fibromuscular dysplasia (FMD) in patients with acute type A aortic dissection (ATAAD). Between 2007 and 2024, 214 patients aged 18–70 years who underwent emergency surgery for ATAAD at Erasmus Medical Center were screened. Fourteen patients were excluded due to the absence of diagnostic CTA imaging, leaving 200 patients for analysis. CTA examinations were evaluated for imaging signs consistent with FMD. Cases with suspected FMD were independently reviewed by a second experienced radiologist, and the final diagnosis of FMD was established by consensus.

Separate carotid CTA examinations were available in 49/200 patients (25%), including 7/18 patients with FMD (39%). Among these seven patients, three demonstrated imaging features consistent with FMD in the carotid arteries.

### Clinical characteristics and sex differences

Of the 18 patients with FMD, 12 were female (Table 1). The proportion of FMD was higher in female patients (12/62, 19.4%; 95% CI, 11.4–30.9%) compared with male patients (6/138, 4.3%; 95% CI, 2.0–9.2%; P<0.001) (Figure 2). Comparison of clinical characteristics between patients with and without FMD revealed no significant differences in age, history of hypertension, or current smoking status.

**Figure 2:**
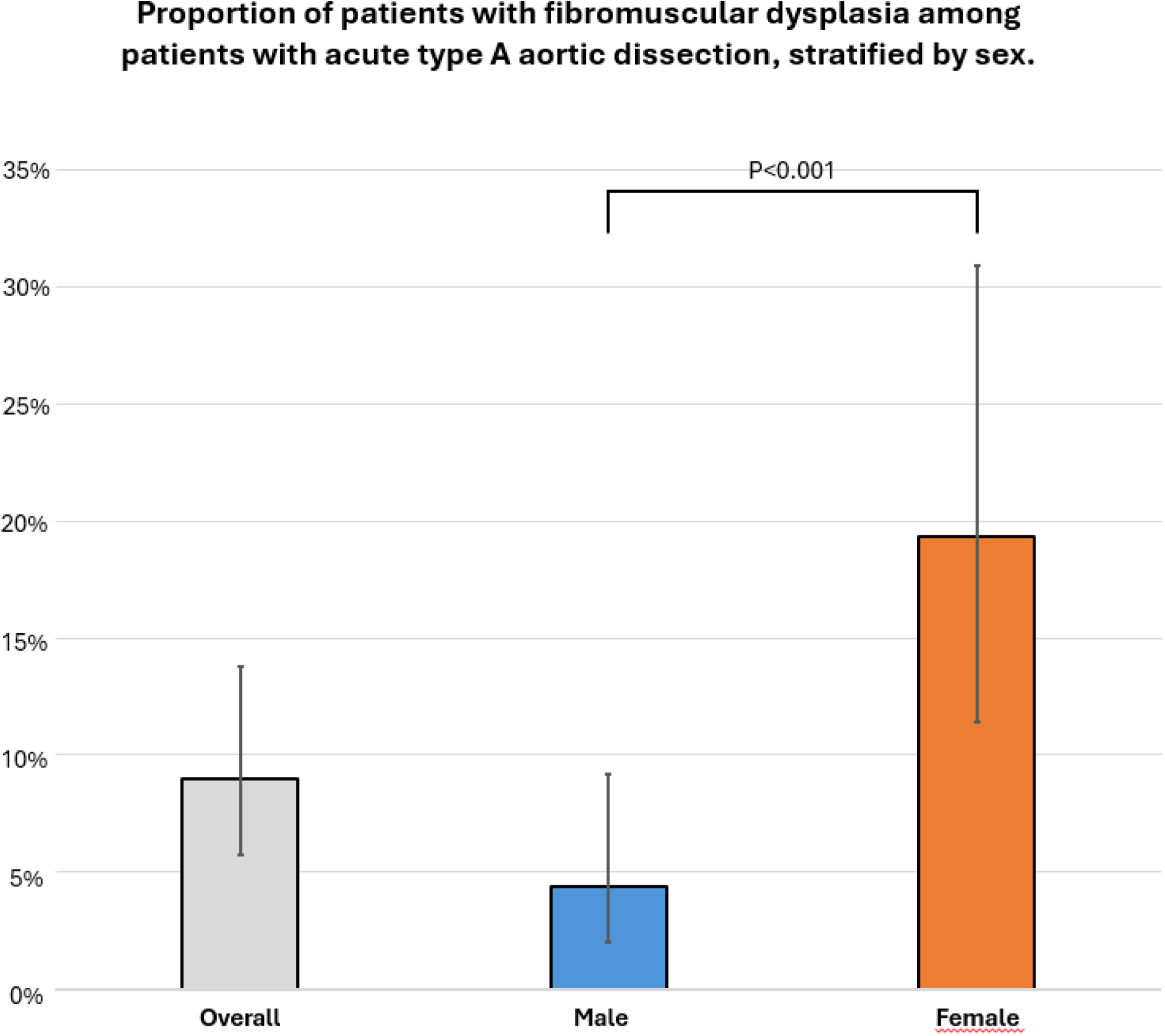
Proportion of patients with fibromuscular dysplasia among patients with acute type A aortic dissection (ATAAD), stratified by sex. A total of 200 patients with ATAAD were included, consisting of 138 male patients and 62 female patients. Error bars indicate 95% confidence intervals.

**Table 1:** Baseline characteristics of patients with acute type A aortic dissection according to fibromuscular dysplasia status.

|  | **Overall (n=200)** | **No FMD (n=182)** | **FMD (n=18)** | **P-value** |
|---|---|---|---|---|
| **Age** | 56.5 ± 9 | 56.2 ± 9 | 59.3 ± 8 | 0.17 |
| **Female** | 62 (31%) | 50 (28%) | 12 (67%) | <0.001 |
| **History of hypertension** | 92 (46%) | 83 (46%) | 9 (50%) | 0.72 |
| **Diabetes** | 1 (0.5%) | 1 (0.5%) | 0 (0%) | 1.00 |
| **COPD** | 8 (4%) | 7 (4%) | 1 (6%) | 1.00 |
| **Current smoking (unknown 18)\*** | 52 (29%) | 46 (28%) | 6 (35%) | 0.52 |
| **Prior aortic surgery** | 5 (3%) | 5 (3%) | 0 (0%) | 1.00 |
| **Prior myocardial infarction** | 8 (4%) | 8 (4%) | 0 (0%) | 1.00 |
| **Prior ischemic cerebrovascular event** | 4 (2%) | 4 (2%) | 0 (0%) | 1.00 |
| **Family history of aortic dissection (unknown 50)\*** | 51 (34%) | 47 (35%) | 4 (29%) | 1.00 |
| **Family history of connective tissue disease (unknown 7)\*** | 31 (16%) | 29 (16%) | 2 (12%) | 0.75 |
| **Genetically confirmed or suspected connective tissue disease (unknown 2)\*** † | 19 (10%) | 17 (9%) | 2 (11%) | 0.68 |

\* Percentages were calculated using available data; patients with missing values were excluded from the denominator.
† Genetically confirmed connective tissue disease included Marfan syndrome (n=4), Loeys–Dietz syndrome (n=2), SMAD3-related disease (n=3), Turner syndrome (n=1), ACTA2-related disease (n=1), and MYH11-related disease (n=1). Seven additional patients had connective tissue disease not otherwise specified.

Baseline characteristics stratified by sex are shown in Table 2. Female patients more frequently had genetically confirmed or suspected connective tissue disease than male patients (10/62, 16% vs. 9/138, 7%, *P*=0.032). Genetically confirmed connective tissue disease was present in 12 patients, including Marfan syndrome (n=4), Loeys-Dietz syndrome (n=2), SMAD3-related disease (n=3), Turner syndrome (n=1), ACTA2 mutation (n=1), and MYH11 mutation (n=1). An additional seven patients had suspected connective tissue disease without a confirmed genetic diagnosis. No other significant sex-related differences in baseline characteristics were observed.

**Table 2:**
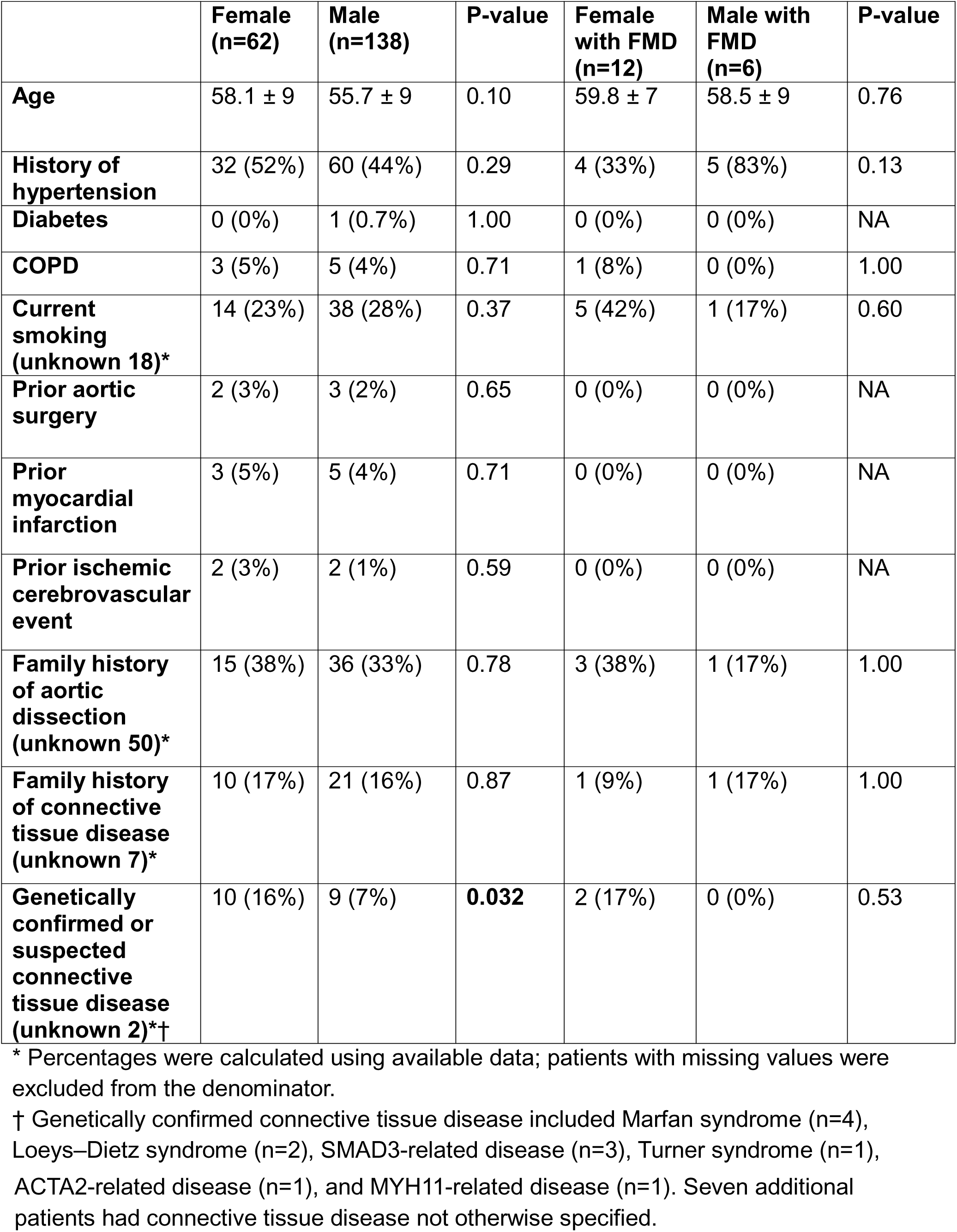
Baseline characteristics stratified by sex in the overall cohort and in the fibromuscular dysplasia subgroup.

### Imaging characteristics of FMD

Representative examples of renal artery FMD in patients with ATAAD are shown in Figure 3.

**Figure 3:**
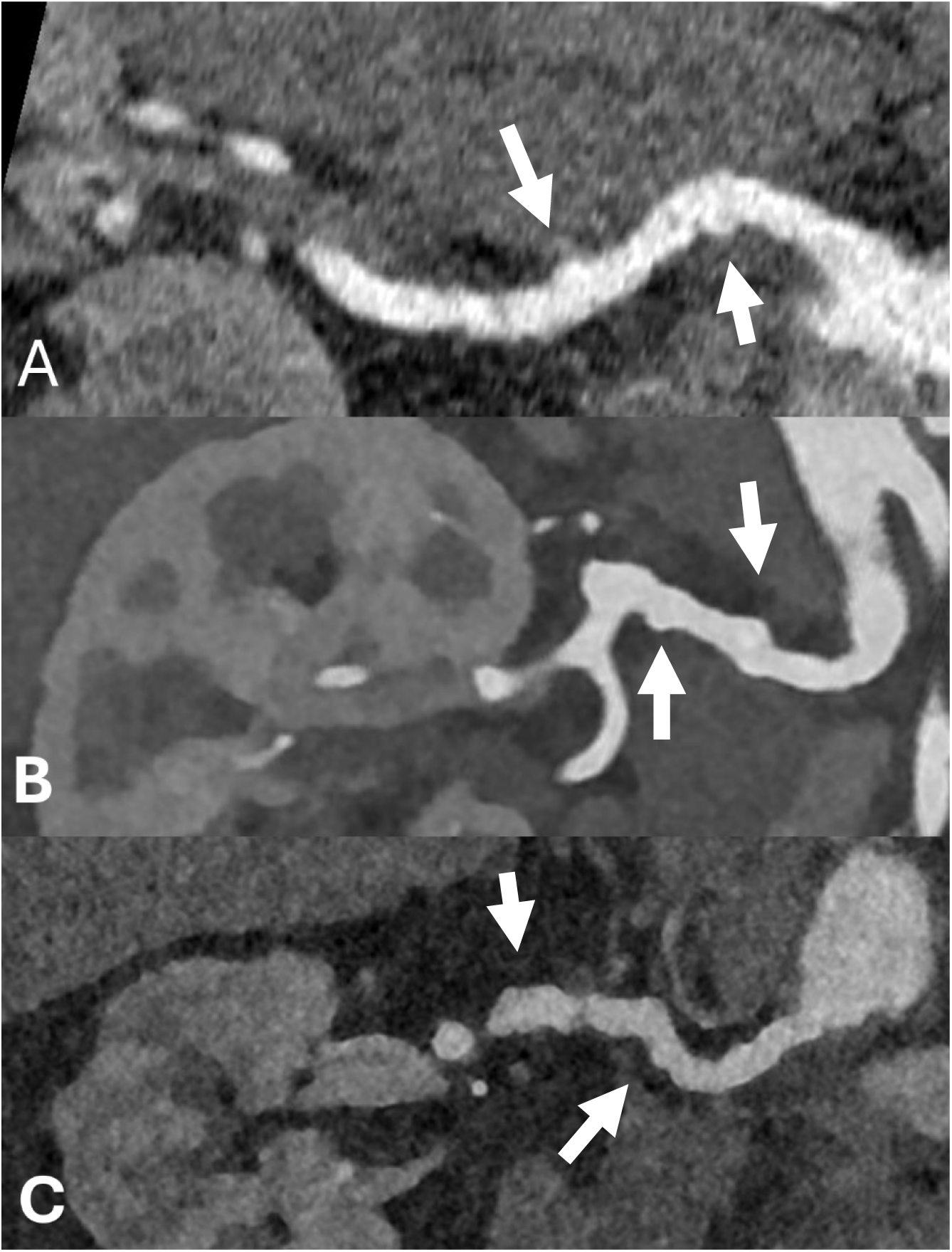
Three case examples of fibromuscular dysplasia involving the right renal artery. Representative curved multiplanar reformations (MPRs) from CTA demonstrating multifocal beading of the mid segment of the right renal artery in patients with ATAAD and FMD. Panel A shows the findings in a woman aged 66–70 years, panel B in a woman aged 61–65 years, and panel C in a man aged 41–45 years. Arrows indicate example locations of characteristic multifocal beading of the renal artery.

Detailed clinical and imaging characteristics of the 18 patients with FMD are presented in Table 3. Within the FMD group, two patients were identified with an underlying connective tissue disease. One patient was a woman aged 56–60 years with Marfan syndrome, demonstrating moderate beading of the right renal artery and typical beading of the internal carotid arteries, findings that are not typically associated with Marfan syndrome. The second patient was a woman aged 61–65 years with ATAAD carrying pathogenic variants in the COL3A1 and SMAD6 genes. CTA demonstrated typical beading of the right renal artery, discrete beading of the right external iliac artery, typical beading with an aneurysm of the left external iliac artery, and typical beading of both distal extracranial internal carotid arteries (Figure 4).

**Figure 4:**
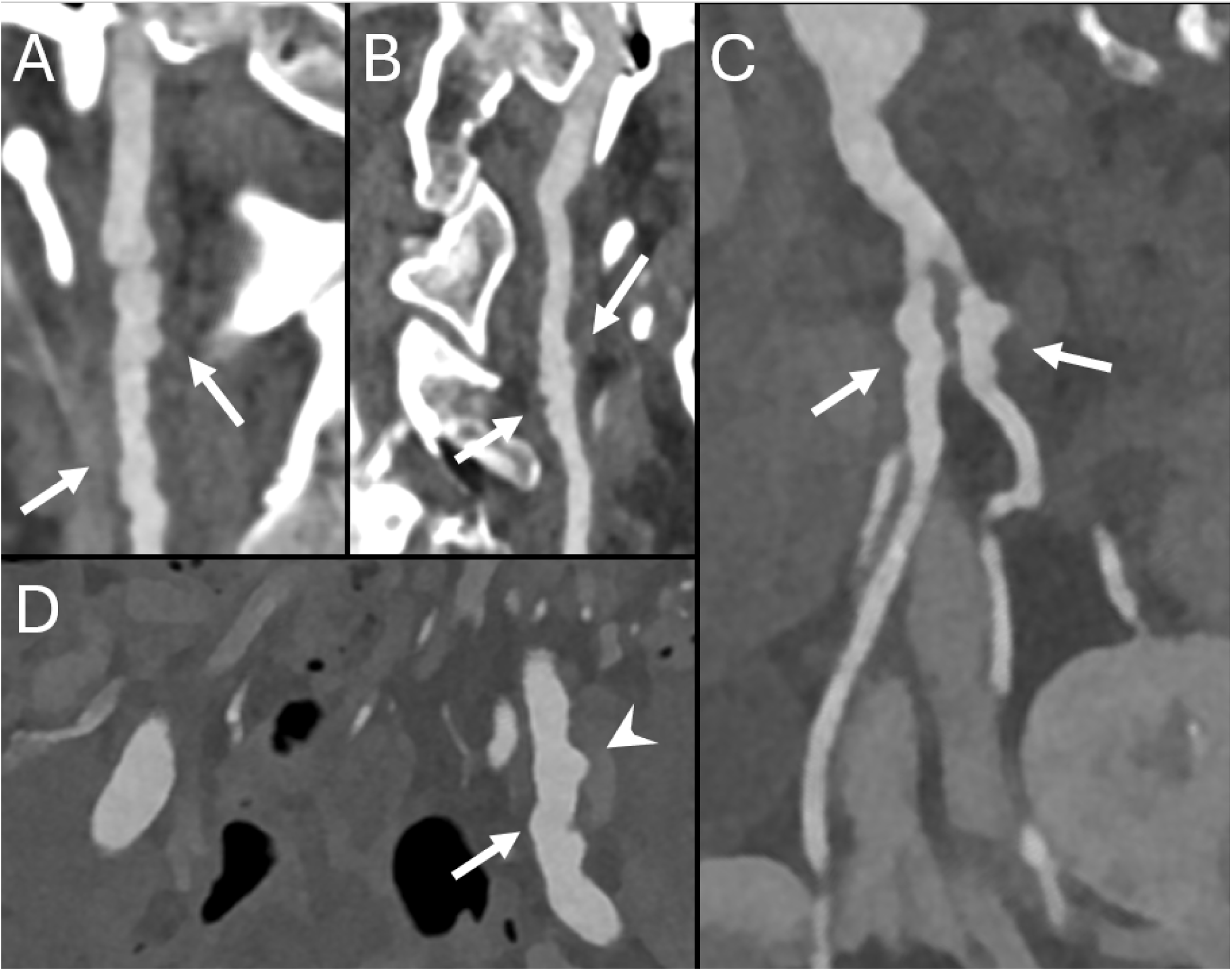
Case example of FMD involving multiple vascular territories. Multivessel fibromuscular dysplasia in a woman aged 61–65 with acute type A aortic dissection and pathogenic variants in the COL3A1 and SMAD6 genes. Curved multiplanar reformations (MPRs) demonstrate multifocal beading of the right internal carotid artery (A), left internal carotid artery (B), right renal artery (C), and left external iliac artery (D) (arrows). An aneurysm of the left external iliac artery is also present (arrowhead in D).

**Table 3:**
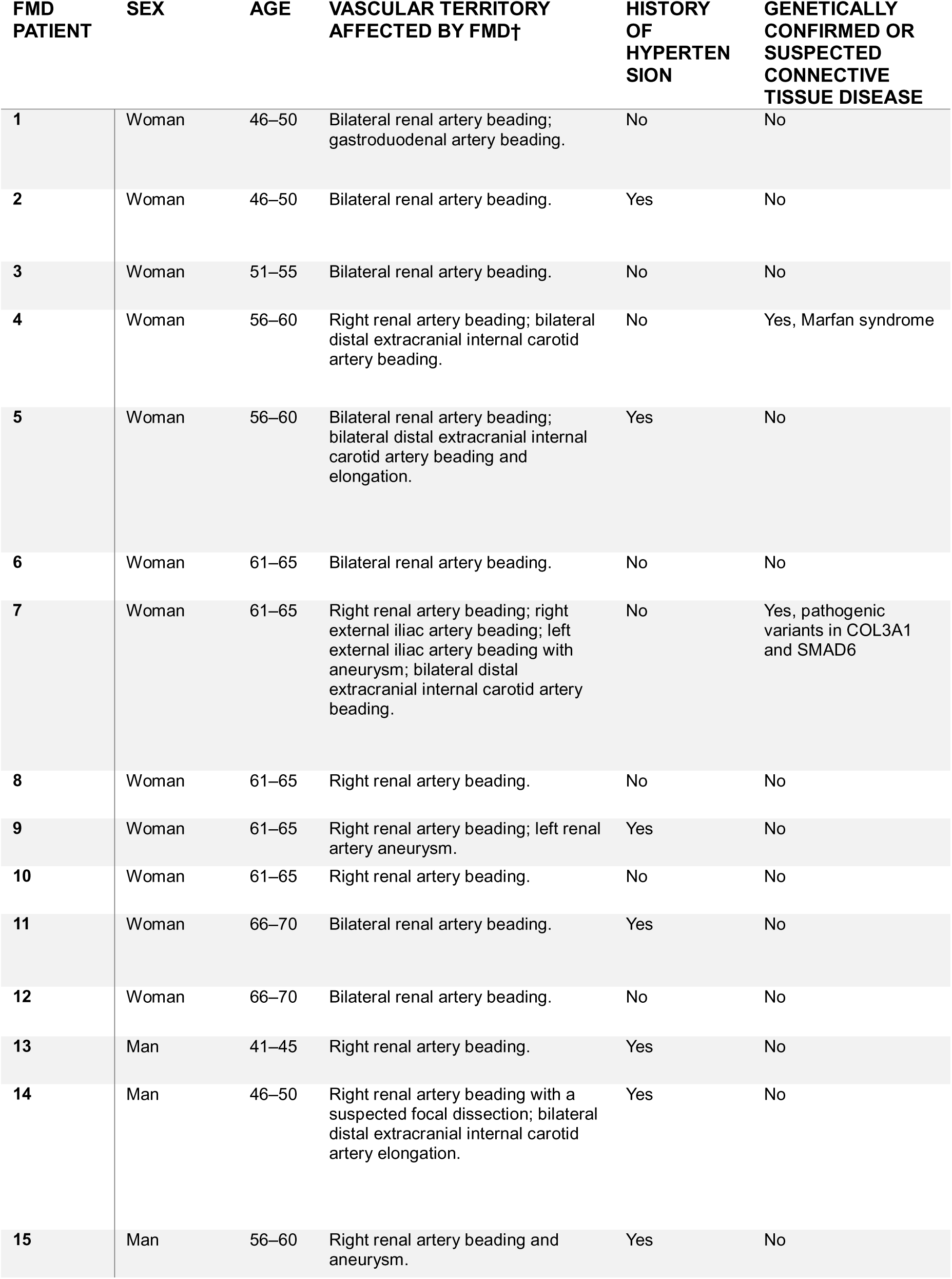
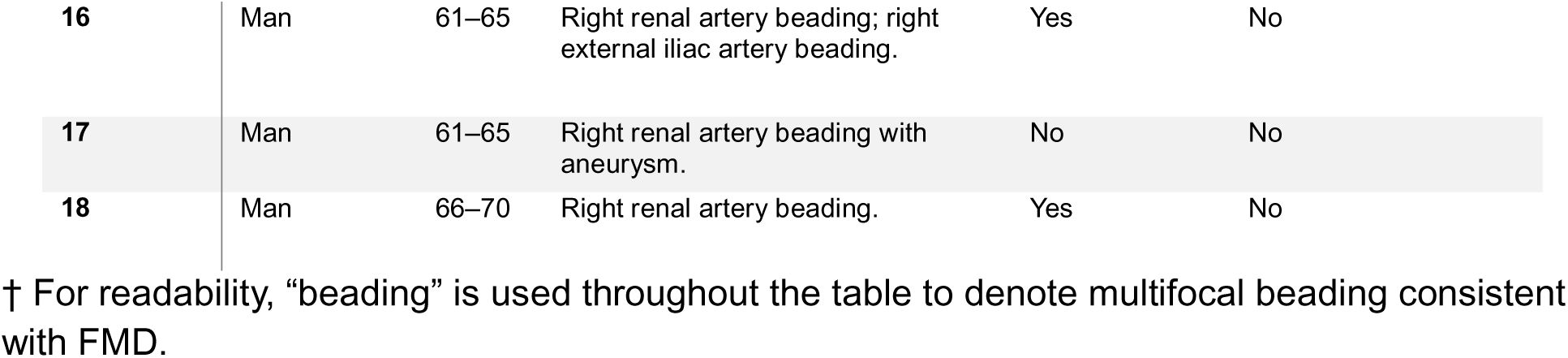
Clinical and imaging characteristics of patients with fibromuscular dysplasia.

| **FMD PATIENT** | **SEX** | **AGE** | **VASCULAR TERRITORY AFFECTED BY FMD†** | **HISTORY OF HYPERTENSION** | **GENETICALLY CONFIRMED OR SUSPECTED CONNECTIVE TISSUE DISEASE** |
|---|---|---|---|---|---|
| **1** | Woman | 46–50 | Bilateral renal artery beading; gastroduodenal artery beading. | No | No |
| **2** | Woman | 46–50 | Bilateral renal artery beading. | Yes | No |
| **3** | Woman | 51–55 | Bilateral renal artery beading. | No | No |
| **4** | Woman | 56–60 | Right renal artery beading; bilateral distal extracranial internal carotid artery beading. | No | Yes, Marfan syndrome |
| **5** | Woman | 56–60 | Bilateral renal artery beading; bilateral distal extracranial internal carotid artery beading and elongation. | Yes | No |
| **6** | Woman | 61–65 | Bilateral renal artery beading. | No | No |
| **7** | Woman | 61–65 | Right renal artery beading; right external iliac artery beading; left external iliac artery beading with aneurysm; bilateral distal extracranial internal carotid artery beading. | No | Yes, pathogenic variants in COL3A1 and SMAD6 |
| **8** | Woman | 61–65 | Right renal artery beading. | No | No |
| **9** | Woman | 61–65 | Right renal artery beading; left renal artery aneurysm. | Yes | No |
| **10** | Woman | 61–65 | Right renal artery beading. | No | No |
| **11** | Woman | 66–70 | Bilateral renal artery beading. | Yes | No |
| **12** | Woman | 66–70 | Bilateral renal artery beading. | No | No |
| **13** | Man | 41–45 | Right renal artery beading. | Yes | No |
| **14** | Man | 46–50 | Right renal artery beading with a suspected focal dissection; bilateral distal extracranial internal carotid artery elongation. | Yes | No |
| **15** | Man | 56–60 | Right renal artery beading and aneurysm. | Yes | No |
| **16** | Man | 61–65 | Right renal artery beading; right external iliac artery beading. | Yes | No |
| **17** | Man | 61–65 | Right renal artery beading with aneurysm. | No | No |
| **18** | Man | 66–70 | Right renal artery beading. | Yes | No |

† For readability, “beading” is used throughout the table to denote multifocal beading consistent with FMD.

## Discussion

In this retrospective study of patients aged ≤70 years, FMD was present in 9% of patients with ATAAD. The proportion of patients with FMD was significantly higher among women (19%) than among men (4%). These findings suggest that FMD may be an underrecognized vascular phenotype associated with ATAAD, particularly in women, and highlight the importance of increased awareness of FMD in this population.

The proportion of patients with FMD in our cohort (9% overall and 19% among women) was higher than the previously reported prevalence estimates of FMD (approximately 3%) in potential kidney donors.[11–13] However, the approximately 4.5-fold higher proportion of FMD in women compared with men in our cohort was consistent with the female predominance observed in potential kidney donor populations, in which FMD was approximately 3.3-fold more prevalent among women.[11] A higher prevalence of FMD has also been reported in older patients with hypertension and a higher atherosclerotic disease burden. In a substudy of the CORAL trial, which investigated renal artery stenting in patients with hypertension, FMD was an exclusion criterion, yet core laboratory analysis identified FMD in 5.8% of patients overall and in 8.8% of women.[14]

Advances in CT technology and improved spatial resolution may also have contributed to increased detection of subtle multifocal arterial beading compared with earlier studies.[19]

This study was not designed to determine whether FMD is causally related to ATAAD. Hypertension associated with FMD could potentially contribute to the risk of ATAAD, as hypertension, particularly when untreated, is a well-established risk factor for ATAAD.[20] However, no significant difference in the proportion of patients with known hypertension was observed between patients with and without FMD in our cohort.

Whether structural alterations of the aortic wall in patients with FMD may independently increase susceptibility to dissection remains unclear. Schwartz et al. reported slightly larger aortic diameters in patients with FMD compared with healthy controls.[15] However, it is uncertain whether this represents direct aortic involvement in FMD or reflects associated factors such as hypertension.

Identifying FMD in patients after ATAAD may be clinically relevant, as it may prompt assessment of associated hypertension and guide additional vascular imaging. Current international guidelines recommend screening of the carotid and intracranial arteries at least once after a diagnosis of FMD.[8]

Radiologists are well positioned to recognize previously undiagnosed FMD in patients with ATAAD, as CTA is routinely performed as part of the diagnostic workup. Careful assessment of the visualized branch vessels may reveal characteristic imaging findings of FMD that might otherwise remain unrecognized. In our cohort, none of the patients with FMD had a previously established diagnosis.

In the overall cohort, approximately 10% of patients had genetically confirmed or suspected connective tissue disease. Among patients with FMD, two individuals had a known genetic disorder: one with Marfan syndrome and one carrying pathogenic variants in the COL3A1 and SMAD6 genes. Both patients demonstrated typical beading of the distal internal carotid arteries. Although this observation may initially appear counterintuitive, it may reflect a more heterogeneous disease spectrum, in which patients can harbor multiple underlying conditions contributing to aortic vasculopathy.

The genetic basis of FMD remains incompletely understood. In previous studies investigating FMD patient cohorts and genes associated with genetic aortic disease (including COL3A1, ACTA2, and TGF-β pathway genes), no relevant contributing genetic variants were identified.[10; 21] Future studies investigating genetic predisposition in patients with both FMD and ATAAD may provide further insight into potential shared or overlapping genetic mechanisms.

Sex-related differences have also been described in patients with thoracic aortic disease. Women tend to be older at the time of diagnosis and more frequently present with tubular ascending aortic aneurysms, whereas men more often exhibit aortic root involvement.[18] The marked female predominance of FMD observed in our ATAAD cohort may represent an additional sex-related difference in the vascular phenotype of these patients.

This study has several limitations. First, this was a retrospective study performed in a single tertiary center, which may have introduced referral center bias and limited generalizability to other populations, although this was partly mitigated by the consecutive inclusion of all eligible surgically treated patients aged 18–70 years during the study period. Second, CTA examinations were performed as part of routine clinical care and were not specifically optimized for the detection of FMD, which may have resulted in underdetection of subtle imaging features of FMD. In addition, second-reader assessment was limited to cases suspected of FMD during the initial review; therefore, subtle findings not identified during the initial review would not have undergone independent second-reader assessment. Furthermore, image assessment was not blinded to patient sex, which may have introduced observer bias in the detection of FMD. However, complete blinding to sex was not feasible because sex-related anatomical characteristics were visible on the CTA examinations. Third, imaging of vascular territories outside the standard aortic CTA scan range, particularly the carotid arteries, was not available in all patients, which may have influenced the detection of FMD. Because carotid CTA was available in only 25% of patients, the proportion of patients with FMD may have been underestimated. Fourth, the relatively small number of patients with confirmed FMD limits the ability to perform more detailed subgroup analyses and may restrict the interpretation of associations with clinical risk factors.

Finally, no systematic genetic testing was performed specifically as part of this study, which limits the ability to explore potential genetic relationships between FMD and ATAAD.

## Conclusion

The current findings suggest that FMD may represent a previously underrecognized vascular phenotype in ATAAD, particularly among women. The presence of patients with both FMD and genetically confirmed or suspected connective tissue disease further highlights the heterogeneity of this patient population. Radiologists play an important role in identifying previously unrecognized FMD through careful assessment of the branch vessels on CTA in patients with ATAAD. Future studies should explore the genetic and vascular characteristics of patients with both FMD and ATAAD to better understand potential shared disease pathways and their relevance to the risk of ATAAD.

## Data Availability

All data produced in the present study are available upon reasonable request to the authors

## Acknowledgments

During manuscript preparation, the authors used ChatGPT (OpenAI) for editorial assistance, including language editing and improving textual consistency. All changes were reviewed and approved by the authors, who take full responsibility for the content of the manuscript.

## REFERENCES

1 Clouse WD, Hallett JW, Jr., Schaff HV (2004) Acute aortic dissection: population-based incidence compared with degenerative aortic aneurysm rupture. Mayo Clin Proc 79:176–180

2 Howard DPJ, Banerjee A, Fairhead JF, et al. (2013) Population-based study of incidence and outcome of acute aortic dissection and premorbid risk factor control: 10-year results from the Oxford Vascular Study. Circulation 127:2031–2037

3 Pape LA, Awais M, Woznicki EM, et al. (2015) Presentation, diagnosis, and outcomes of acute aortic dissection: 17-year trends from the International Registry of Acute Aortic Dissection (IRAD). J Am Coll Cardiol 66:350–358

4 Evangelista A, Isselbacher EM, Bossone E, et al. (2018) Insights from the International Registry of Acute Aortic Dissection: a 20-year experience of collaborative clinical research. Circulation 137:1846–1860

5 Czerny M, Schmidli J, Adler S, et al. (2019) Current options and recommendations for the treatment of thoracic aortic pathologies involving the aortic arch: an expert consensus document of the European Association for Cardio-Thoracic Surgery (EACTS) and the European Society for Vascular Surgery (ESVS). Eur J Cardiothorac Surg 55:133–162

6 Nienaber CA, Clough RE, Sakalihasan N et al (2016) Aortic dissection. Nat Rev Dis Primers 2:16053

7 Pape LA, Tsai TT, Isselbacher EM, et al. (2004) Aortic dissection in women: findings from the International Registry of Acute Aortic Dissection. J Am Coll Cardiol 43:665–671

8 Gornik HL, Persu A, Adlam D et al (2019) First International Consensus on the diagnosis and management of fibromuscular dysplasia. Vasc Med 24:164–189

9 Kadian-Dodov D, Gornik HL, Gu X et al (2016) Dissection and Aneurysm in Patients With Fibromuscular Dysplasia: Findings From the U.S. Registry for FMD. J Am Coll Cardiol 68:176–185

10 Olin JW, Gornik HL, Bacharach JM et al (2014) Fibromuscular dysplasia: state of the science and critical unanswered questions: a scientific statement from the American Heart Association. Circulation 129:1048–1078

11 Lorenz EC, Vrtiska TJ, Lieske JC et al (2010) Prevalence of renal artery and kidney abnormalities by computed tomography among healthy adults. Clin J Am Soc Nephrol 5:431–438

12 McKenzie GA, Oderich GS, Kawashima A, Misra S (2013) Renal artery fibromuscular dysplasia in 2,640 renal donor subjects: a CT angiography analysis. J Vasc Interv Radiol 24:1477–1480

13. Plouin PF, Perdu J, La Batide-Alanore A, Boutouyrie P, Gimenez-Roqueplo AP, Jeunemaitre X (2007) Fibromuscular dysplasia. Orphanet J Rare Dis 2:28

14 Hendricks NJ, Matsumoto AH, Angle JF et al (2014) Is fibromuscular dysplasia underdiagnosed? A comparison of the prevalence of FMD seen in CORAL trial participants versus a single institution population of renal donor candidates. Vasc Med 19:363–367

15 Schwartz AM, Kim E, Gleason P, Li X, Ko YA, Wells BJ (2022) Aortic Dimensions Are Larger in Patients With Fibromuscular Dysplasia. J Am Heart Assoc 11:e023858

16 Tasaki T, Hatanaka K, Kirishima M, et al. (2017) Aortic fibromuscular dysplasia complicated by dissection: a case report and review of literature. Cardiovasc Pathol 31:41–46

17 Kar S, Gopaldas RR, Kumar A (2013) Acute aortic dissection and stroke in multivessel fibromuscular dysplasia. Tex Heart Inst J 40:88–90

18. Notenboom ML, de Keijzer AR, Veen KM, et al (2025) Sex-related differences in the clinical course of aortic root and ascending aortic aneurysms: the DisSEXion Study. Eur Heart J 46:551–564

19 Greffier J, Viry A, Robert A, Khorsi M, Si-Mohamed S (2025) Photon-counting CT systems: A technical review of current clinical possibilities. Diagnostic and Interventional Imaging 106:53–59

20 Hibino M, Otaki Y, Kobeissi E et al (2022) Blood Pressure, Hypertension, and the Risk of Aortic Dissection Incidence and Mortality: Results From the J-SCH Study, the UK Biobank Study, and a Meta-Analysis of Cohort Studies. Circulation 145:633–644

21 Poloskey SL, Kim E, Sanghani R et al (2012) Low yield of genetic testing for known vascular connective tissue disorders in patients with fibromuscular dysplasia. Vasc Med 17:371–378

